# Reliability of a Simple, Biomechanically Grounded Score for Landing-Mechanics Evaluation

**DOI:** 10.64898/2026.02.09.26345781

**Authors:** Shinsuke Sakoda, Kouichi Kajiwara, Rikiya Shito, Hiroto Kumagae, Osamu Yokoi, Kimiaki Kawano

## Abstract

**Context:** Clinical assessments of landing mechanics often require complex scoring systems or laboratory-based motion analysis, which can limit feasibility in routine practice. A visually based landing-mechanics score centered on a standardized optimal joint-alignment configuration (“Zero Position”) may offer a simple, clinically deployable alternative.

**Objective:** To determine the intra- and inter-rater reliability of a landing mechanics score based on standardized optimal joint alignment at the moment of maximal center-of-mass (COM) descent.

**Design:** Cross-sectional reliability study.

**Setting:** University athletic training facility.

**Patients or Other Participants:** Ninety healthy male collegiate athletes.

**Main Outcome Measures:** Landing mechanics were evaluated using frontal- and sagittal-plane video recordings, with scoring performed on the frame corresponding to maximal COM descent. Five criteria reflecting the standardized joint configuration (“Zero Position”) were assessed. Intra- and inter-rater reliability were calculated using Cohen’s kappa coefficients and Kendall’s W.

**Results:** All five criteria demonstrated moderate to substantial intra-rater reliability and moderate to almost perfect inter-rater reliability. The total landing-mechanics score showed excellent agreement across all comparisons. The scoring system required minimal training and was feasible to implement using standard video recordings.

**Conclusions:** The landing-mechanics score centered on the Zero Position demonstrated high reliability and strong clinical feasibility. This simple, visually grounded assessment may support routine clinical screening, injury-risk evaluation, and return-to-sport decision-making. Future research should examine its applicability to single-leg landings and sport-specific high-risk movements.

## Introduction

Non-contact lower extremity injuries, including anterior cruciate ligament (ACL) tears and lateral ankle sprains, frequently occur during dynamic athletic tasks such as landing, cutting, and rapid deceleration. Epidemiologic and biomechanical studies indicate that these injuries often occur within the first 50–100 milliseconds after ground contact and are associated with movement patterns such as dynamic knee valgus, insufficient hip and knee flexion, and poor trunk control. ^1,2^ Early identification of these high-risk mechanics is therefore essential for effective injury prevention and rehabilitation.

Although three-dimensional motion analysis has substantially advanced our understanding of landing biomechanics, its high cost, technical demands, and limited clinical practicality restrict its routine use. ^3, 4^ Methodological issues—including marker placement error, soft-tissue artifact, and variability in joint-angle estimation—further limit its feasibility for widespread screening applications. ^5, 6^

Simplified field-applicable tools such as the Landing Error Scoring System (LESS) and the Drop Jump Screening Test (DJST) have been developed as alternatives. However, the LESS includes numerous scoring items that may reduce consistency, whereas the DJST requires specialized equipment and software, limiting practicality in many clinical and field environments. ^7, 8^

To address these limitations, we developed a clinically practical landing evaluation score centered on a single biomechanically critical moment: the instant of maximal center-of-mass (COM) descent. This phase corresponds to a period of high mechanical load and maximal internal joint-moment demand, making it an ideal time point for evaluating shock-absorption strategies and lower-extremity alignment.”

At this moment, a favorable posture―historically and biomechanically conceptualized as a balanced flexed-limb position―typically involves approximately 90° of hip, knee, and ankle flexion, which provides a mechanically advantageous configuration for force attenuation and dynamic stability. We refer to this optimal joint configuration as the “Zero Position.” This configuration also reflects a balanced, mechanically advantageous posture that is commonly adopted as athletes prepare for subsequent movements, underscoring its functional relevance beyond the landing phase.

Therefore, the purpose of this study was to introduce a simplified landing mechanics score based on this optimal Zero Position alignment and to evaluate its intra- and inter-rater reliability.

## Methods

### Participants

Ninety healthy male collegiate athletes (all aged 18 years at the time of testing, assessed at university entry) participated in this study.

### Landing Task

Participants performed a standardized drop-landing task based on the protocol described in the Drop Jump Screening Test (DJST) by Noyes et al. ^7^ Participants stepped off a 30-cm platform, landed bilaterally, and then performed a maximal vertical jump following the established procedure.

Only the initial landing phase was analyzed. For each trial, the frame corresponding to the instant of maximal center-of-mass (COM) descent—defined as the lowest vertical position of the trunk midpoint, used as a practical surrogate for the COM—was selected for scoring.

### Video Recording

Frontal- and sagittal-plane video recordings were obtained using a single digital camera (iPhone; Apple Inc).

The camera was positioned approximately 3 m from the landing area and mounted on a tripod at the height of the participant’s greater trochanter to minimize parallax error. Videos were recorded at 120 frames per second under adequate lighting to ensure clear visualization of joint landmarks.

### Landing Mechanics Scoring Criteria

Five scoring criteria were developed to evaluate lower-extremity alignment at the moment of maximal COM descent. The center of mass (COM) was operationally approximated using the midpoint of the trunk as a surrogate marker.

The target joint angles (approximately 90° for hip, knee, and ankle dorsiflexion) were selected based on prior biomechanical literature describing optimal muscle length– tension relationships, ^12^ maximal moment-arm mechanics in flexed postures, ^13^ and clinically observed shock-absorption strategies reported in foundational gait and sports-movement analyses. ^10–11^ These definitions were finalized by consensus among experienced sports-medicine clinicians.

### Sagittal-plane criteria

1. Knee Flexion Angle — approximately 90°.
2. Ankle Dorsiflexion Angle — angle formed by the MTP–malleolus segment and the tibia; approximately 90°.
3. Hip Flexion / Trunk Forward Lean — approximately 90°, with the trunk positioned nearly parallel to the tibia.

#### Frontal-plane criterion

4. Knee Alignment — no dynamic valgus or varus.

#### Combined criterion

5. COM Vertical Stability — COM maintained over the base of support (BOS). The BOS was referenced to medial forefoot, reflecting the primary point of load acceptance during landing.

Although the present study focused on bilateral landings, the conceptual framework and scoring criteria are applicable to single-leg landings, which will be evaluated in future investigations.

### Raters and Training

Raters were sports medicine clinicians with at least 3 years of clinical experience. All raters completed standardized training that included familiarization with the scoring criteria and exemplar video cases.

### Scoring Procedure

All videos were anonymized prior to scoring. To evaluate intra-rater reliability, each rater scored all trials twice with a 2-week washout period between sessions. Raters were blinded to their previous ratings and to each other’s evaluations.

### Analysis

Cohen’s kappa coefficients were calculated for each of the five scoring criteria to assess intra- and inter-rater reliability. Kendall’s W was used to assess agreement for the total landing-mechanics score.

### Ethical approval

This study involved non-invasive video analysis of healthy adult athletes performing a standardized landing task. All participants provided informed consent prior to participation. According to institutional policy, formal ethical committee approval was not required for this type of minimal-risk observational study.

## Results

A total of 90 landing trials were analyzed. All five landing-mechanics criteria demonstrated moderate to substantial intra-rater reliability and moderate to almost perfect inter-rater reliability, as summarized in Table 1.

**Table 1.** Intra-and inter-rater reliability for the five landing-mechanics criteria (Cohen’s kappa coefficients).

| Criterion | Intra-rater Reliability |  | Inter-rater Reliability |  |
| --- | --- | --- | --- | --- |
|  | ExaminerA | ExaminerB | 1st session | 2nd session |
| 1 | 0.63 (0.45-0.81) | 0.54 (0.33-0.74) | 0.73 (0.57-0.90) | 0.82 (0.69-0.95) |
| 2 | 0.66 (0.51-0.81) | 0.66 (0.51-0.82) | 0.75 (0.61-0.89) | 0.84 (0.73-0.95) |
| 3 | 0.77 (0.63-0.90) | 0.60 (0.42-0.77) | 0.72 (0.58-0.87) | 0.68 (0.52-0.84) |
| 4 | 0.42 (0.22-0.63) | 0.53 (0.35-0.71) | 0.63 (0.46-0.79) | 0.76 (0.62-0.91) |
| 5 | 0.62 (0.45-0.80) | 0.63 (0.45-0.80) | 0.72 (0.56-0.87) | 0.83 (0.70-0.95) |

Values represent Cohen's kappa coefficients with 95% confidence intervals. Criteria 1–5 correspond to the five landing-mechanics scoring items.

For intra-rater reliability, Cohen’s kappa coefficients ranged from 0.42 to 0.77 for Examiner A and from 0.53 to 0.66 for Examiner B. For inter-rater reliability, kappa coefficients ranged from 0.63 to 0.84, indicating consistently strong agreement between evaluators across all criteria. The total landing-mechanics score demonstrated excellent agreement among all comparisons (Table 2). Kendall’s W values were 0.88 for Examiner A (first and second sessions; both p < 0.001), 0.88 for Examiner B (first and second sessions; both p < 0.001), and ranged from 0.91 to 0.93 for inter-rater comparisons (both p < 0.001).

**Table 2.** Intra- and inter-rater reliability for the total landing-mechanics score (Kendall’s W).

| Intra-rater Reliability |  | Inter-rater Reliability |  |
| --- | --- | --- | --- |
| ExaminerA | ExaminerB | 1st session | 2nd session |
| 0.88 (p<0.001) | 0.88 (p<0.001) | 0.91 (p<0.001) | 0.93 (p<0.001) |

Values represent Kendall's coefficient of concordance (W).

These findings indicate that both individual scoring items and the combined total score can be applied with high consistency following minimal evaluator training.

## Discussion

This study demonstrated that a simple, biomechanically grounded score for landing-mechanics evaluation―focused on the moment of maximal center-of-mass (COM) descent―provides high intra- and inter-rater reliability. By focusing the evaluation on a single biomechanically meaningful frame and reducing the assessment to five essential criteria, the scoring system offers a practical alternative to more complex screening tools traditionally used in sports medicine.

Three-dimensional motion analysis remains the criterion standard for quantifying lower-extremity mechanics; however, its reliance on expensive equipment, marker placement, and technically demanding data processing limits its practicality in most clinical and athletic environments. ^3–6^ Existing field-based alternatives, such as the Landing Error Scoring System (LESS) and the Drop Jump Screening Test (DJST), also present challenges. The LESS includes numerous scoring items that may reduce scoring consistency, while the DJST requires specialized software and equipment not universally available. ^7–8^ In contrast, the present scoring method can be implemented using standard video recordings and minimal evaluator training, suggesting strong feasibility for routine clinical use.

A central feature of this scoring system is the joint configuration that we conceptualized as the “Zero Position.” This posture—characterized by approximately 90° of hip, knee, and ankle flexion—aligns with foundational biomechanical principles describing optimal muscle length–tension relationships^12^ and joint moment-arm mechanics near mid-range flexion. ^13^ At this configuration, major lower-extremity muscle groups operate near their optimal mechanical advantage, facilitating active force attenuation during high-impact tasks.

Non-contact ACL injuries often occur immediately after ground contact and are associated with inadequate lower-extremity alignment and neuromuscular control. ^1, 2^ Achieving the Zero Position at the moment of peak loading may therefore mitigate exposure to hazardous multiplanar knee-loading patterns.

This concept is particularly relevant for sport-related injuries such as ACL rupture, where excessive load is transferred to static stabilizers when dynamic control is insufficient. A landing pattern approximating the Zero Position helps redistribute impact forces away from passive structures and toward the body’s dynamic stabilizers, allowing the musculature to absorb load more effectively and reducing strain on the ACL.

Although achieving a perfectly defined Zero Position during high-speed athletic maneuvers—such as cutting, re-acceleration, or unanticipated directional changes—is often impractical, the underlying concept remains clinically meaningful. Rather than reproducing the exact 90° joint configuration, maintaining movement patterns that approximate the Zero Position—or avoiding substantial deviations from it—may be sufficient to reduce hazardous multiplanar loading. In this sense, the Zero Position serves as a biomechanical reference zone toward which athletes should converge during high-demand tasks, rather than a rigid target posture. This interpretation aligns with injury-mechanism research indicating that large departures from optimal lower-extremity alignment, rather than failure to achieve a fixed ideal angle, are most strongly associated with increased ACL loading.

The Zero Position is also functionally relevant within the broader context of athletic movement. Landing is not simply the termination of a jump but the preparatory position for subsequent actions such as cutting, sprinting, or re-acceleration. We use the term “zero” to denote a biomechanically and functionally balanced baseline that represents not only the end point of a landing movement but also the starting point for the subsequent action.

Although concepts such as the “athletic stance,” “ready position,” or “power position” share superficial similarities, these stances generally lack explicit joint-angle definitions and have been described primarily from a coaching perspective rather than through rigorous biomechanical analysis. In contrast, the Zero Position integrates specific joint-angle criteria derived from established mechanical principles, distinguishing it from traditional preparatory stances.

An important implication of this work is the potential extension of the scoring system to single-leg tasks, which impose greater multiplanar demands and are frequently implicated in ACL injury mechanisms. ^1, 2^ Evaluating whether athletes can maintain optimal alignment during single-leg landings, cutting, or sudden deceleration may enhance both the clinical utility and the injury-prevention relevance of the Zero Position framework. Although the present study focused exclusively on bilateral landings, the conceptual structure of the scoring criteria is inherently applicable to single-leg tasks, supporting future research in this area.

Overall, the findings indicate that the Zero Position–based landing mechanics score provides a reliable and clinically feasible method for assessing lower-extremity alignment during a biomechanically critical phase of landing. Its simplicity, minimal equipment requirement, and strong reliability make it a promising tool for injury-risk screening, rehabilitation monitoring, and return-to-sport decision-making.

### Limitation

Several limitations should be acknowledged. First, the study sample consisted exclusively of healthy, 18-year-old male collegiate athletes, which limits generalizability to female athletes, youth athletes, older individuals, or those with a history of lower-extremity injury. Future studies should examine the applicability and reliability of the scoring system in broader athletic and clinical populations.

Second, all assessments were based on two-dimensional video analysis. Although widely used in clinical practice, 2D imaging cannot fully capture multiplanar motion and may introduce parallax-related error. Validation of the Zero Position using three-dimensional motion analysis would help confirm the biomechanical assumptions underlying the scoring criteria.

Third, raters underwent standardized training, which may have positively influenced scoring consistency. It remains unclear whether clinicians with varying levels of experience—or without structured training—would achieve similar reliability. Evaluating abbreviated or self-directed training protocols would improve the method’s clinical feasibility.

Fourth, the videos were recorded under controlled laboratory conditions with consistent lighting, camera placement, and task execution. Real-world screening environments—including field settings, varied footwear, or unanticipated landings—may produce different results. Field-based testing is needed to evaluate performance under practical conditions.

Finally, the scoring system focused exclusively on bilateral landings and on a single time point: maximal COM descent. Although biomechanically meaningful, landing is a dynamic sequence, and high-risk movements often involve single-leg tasks, cutting, or rapid deceleration. Future research should determine whether maintaining the Zero Position during these more complex tasks predicts injury risk or performance outcomes.

## Conclusion

The landing mechanics score centered on the Zero Position joint configuration demonstrated high intra- and inter-rater reliability and can be efficiently implemented using standard video recordings. Its simplicity, minimal training requirement, and strong consistency support its use in routine clinical and athletic settings. Further validation across diverse populations, single-leg tasks, and sport-specific movements is warranted to establish its broader applicability for injury-risk screening and return-to-sport decision-making.

## Data Availability

All data produced in the present study are available upon reasonable request to the authors

## Notes

### Competing Interest Statement

The authors have declared no competing interest.

### Funding Statement

This study did not receive any funding.

### Author Declarations

The Ethics Committee of Ashiya Central Hospital gave ethical approval for this work.

## References

1. Woods C, Hawkins RD, Maltby S, et al. The Football Association Medical Research Programme: an audit of injuries in professional football—analysis of preseason injuries. Br J Sports Med. 2003;37(5):385–392.

2. Krosshaug T, Nakamae A, Boden BP, et al. Mechanisms of anterior cruciate ligament injury in basketball: video analysis of 39 cases. Am J Sports Med. 2007;35(3):359–367.

3. Hewett TE, Myer GD, Ford KR. Biomechanical measures of neuromuscular control and valgus loading of the knee predict ACL injury risk in female athletes. Am J Sports Med. 2005;33(4):492–501.

4. Sheehan FT, Wilson NA, Paterno MV, et al. A novel method for quantifying lower extremity movement symmetry. Am J Sports Med. 2012;40(2):395–402.

5. Kristianslund E, Krosshaug T, van den Bogert AJ. Effect of low pass filtering on joint moments from inverse dynamics: implications for injury prevention. Sports Biomech. 2012;11(2):127–142.

6. McLean SG, Walker K, Ford KR, et al. Evaluation of a functional preparticipation screening model: assessing the components of the Drop Jump Screening Test. Sports Med. 2005;35(10):813–828.

7. Noyes FR, Barber-Westin SD, Fleckenstein C, et al. The Drop-Jump Screening Test: difference in lower limb control displayed by male and female athletes. Am J Sports Med. 2005;33(2):197–207.

8. Padua DA, Marshall SW, Boling MC, et al. The Landing Error Scoring System (LESS): reliability and factorial validity. Am J Sports Med. 2009;37(10):1996–2002.

9. Iqbal K, Pai YC. Predicted step width and optimal foot placement for the avoidance of a fall in walking. J Biomech. 2000;33(10):1387–1395.

10. Borelli GA. De Motu Animalium. 1680.

11. Perry J, Burnfield JM. Gait Analysis: Normal and Pathological Function. 2nd ed. SLACK; 2010.

12. Lieber RL. Skeletal Muscle: Structure, Function, and Plasticity. 3rd ed. Lippincott Williams & Wilkins; 2010.

13. Krevolin JL, Pandy MG, Pearce JC. Moment arm variations at the knee with changes in joint angle. J Biomech. 2004;37(5):785–795.

14. Behm DG, Chaouachi A. A review of the acute effects of static and dynamic stretching on performance. Eur J Appl Physiol. 2011;111(11):2633–2651.

